# Linking biological variation to outcomes of statin treatment in the general population

**DOI:** 10.1101/2023.03.21.23287344

**Authors:** Iryna Hlushchenko, Mohammad Majharul Islam, Max Tamlander, Samuli Ripatti, Simon George Pfisterer

**Author notes:** **Corresponding author:** Simon Pfisterer Address: Haartmaninkatu 8, Biomedicum 1, Helsinki, 00290, Finland.

## Abstract

Interindividual differences for outcomes of lipid-lowering therapy are well known. Alterations in cellular pathways may contribute to the phenomenon. To address this question, we employed an automated multiplexed analysis pipeline to systematically characterize alterations in cellular lipid trafficking in leukocytes from 400 subjects of the FINRISK 2012 Study. Of individuals receiving high-intensity statin therapy those with lower cellular lipid trafficking scores displayed higher circulating concentrations of several pro-atherogenic lipoproteins and had higher odds for myocardial infarction and stroke when compared to the rest of the subjects with equivalent treatment. Most subjects with a poor lipid trafficking score did not reach low density lipoprotein cholesterol (LDL-C) target levels on statin monotherapy. Lipid trafficking scores showed synergy with a polygenic risk score for LDL-C, improving the association with pro-atherogenic lipoprotein profile when combined. Our results suggest that quantification of cellular lipid trafficking can aid in treatment selection and risk assessment in dyslipidemia.

**Graphical Abstract:** 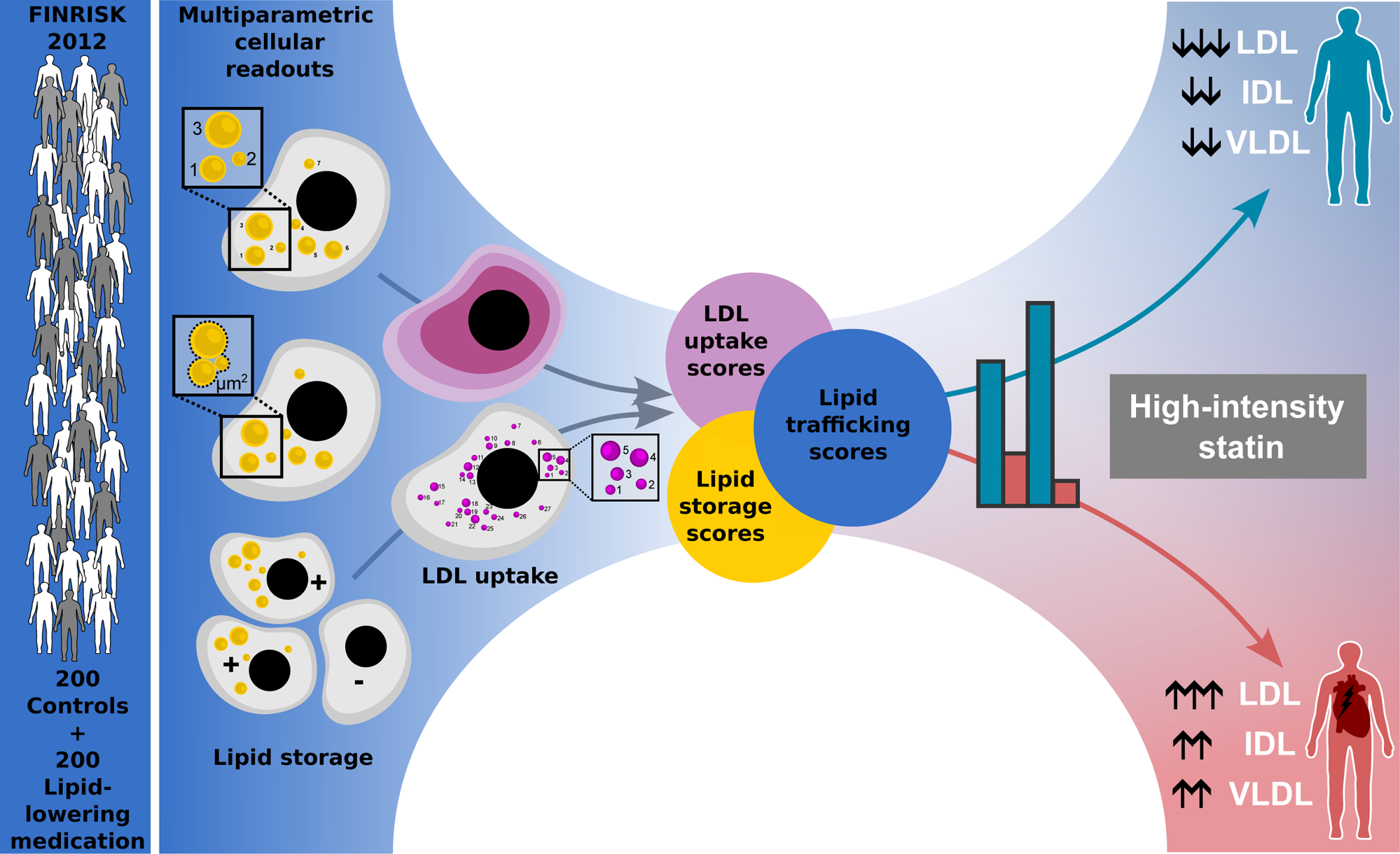

## Introduction

Hypercholesterolemia is a global health problem and the leading cause of cardiovascular disease (CVD).^1^ Even though multiple treatment options exist, such as statins, ezetimibe or PCSK9 inhibitors (PCSK9i), a large fraction of high-risk patients does not achieve desired low-density lipoprotein cholesterol (LDL-C) levels.^2–8^ The reasons for poor goal attainment are multifactorial, including low adherence to the prescribed therapy, as well as challenges imposed through healthcare systems and reimbursement rules.^9–12^ Interindividual variation in treatment outcomes has been reported in several studies,^3,13,14^ including those in which adherence has been monitored closely^15^. Therefore, it appears possible that biological variation in cellular pathways underlying drug action might represent an important contributing factor to goal attainment, but lacks experimental data to receive recognition.

Statins and PCSK9 inhibitors influence cellular lipid trafficking via different mechanisms. Whilst PCSK9i block degradation of the low-density lipoprotein (LDL) receptor (LDLR), statins evoke cellular cholesterol depletion and increased LDLR expression.^16–18^ In both cases more abundant plasma membrane LDLR results in increased LDL uptake and clearance from the blood. Cell surface abundance of LDLR is linked to cellular lipid homeostasis, with excess cholesterol and lipid storage resulting in reduced LDLR expression.^19,20^

For familial hypercholesterolemia (FH) patients, alterations in cellular LDL uptake have been linked to treatment outcomes for statin^21–24^ and PCSK9i medication.^25^ Initially, cellular LDL uptake studies were designed to identify FH patients.^26^ However, differentiation of FH patients from control individuals was difficult, due to interindividual variability in both groups.^27^ Previously, we have established a multiplexed automated analysis pipeline for systematic quantification of cellular LDL uptake and lipid storage in leukocyte subpopulations under different cholesterol depletion states, providing 26 readouts on lipid trafficking for each subject.^21^ We demonstrated that monocytes display more robust responses in both LDL-uptake and lipid storage than lymphocytes and that individuals with identical LDLR mutations display highly divergent LDL uptake profiles which are associated with achieved LDL-C on statin therapy.^21^ This highlights that variations in cellular lipid trafficking pathways are likely widespread, however, it is yet unclear whether they are relevant for lipid-lowering treatment outcomes and cardiovascular disease progression in the general population. Human leukocyte and liver LDLR expression are highly correlated^28^, indicating that leukocyte-derived cellular readouts may be utilized as surrogate readouts for hepatocytes to assess biological variation in the context of dyslipidemia. This is supported by previous studies showing a correlation of leukocyte LDL uptake and circulating LDL-C for FH patients^21,23–25^.

The main objective of this study was to elucidate how LDL-uptake and lipid storage in leukocyte subpopulations associate with circulating LDL-C and different lipoprotein subclasses in statin users from the general population, versus subjects not on lipid-lowering therapy. Secondary goals were to examine if poor lipid trafficking profiles are associated with increased cardiovascular risk and to investigate the interrelationship of cellular and genetic risk scores.

## Results

### Turning multiparametric readouts into cellular lipid trafficking scores

Peripheral blood mononuclear cell (PBMC) samples were analyzed with an automated pipeline for detailed quantification of LDL uptake and lipid storage as described previously. ^21^ In the LDL-uptake assay we quantified the mean intensity of internalized fluorescently labeled LDL (Int) and the number of LDL-filled organelles (No) in lymphocyte and monocyte populations under lipid-rich and -poor conditions (Figure 1A, C). While mean cell intensity reflects the total amount of internalized and surface-bound LDL particles, the number of organelles describes exclusively internalized LDL. In lipid-rich conditions we read out the cellular uptake potential in non-stimulated conditions. When cells are challenged by lipid starvation, LDLR expression is upregulated, increasing LDL uptake from the media. In these conditions we quantify a maximal LDL uptake capacity of a person’s leukocytes. Mean intensities and organelle counts were normalized to controls and combined for each condition into an uptake score for lipid-rich (UPT-R) and lipid-poor conditions (UPT-P) (Figure 1C).

**Figure 1.**
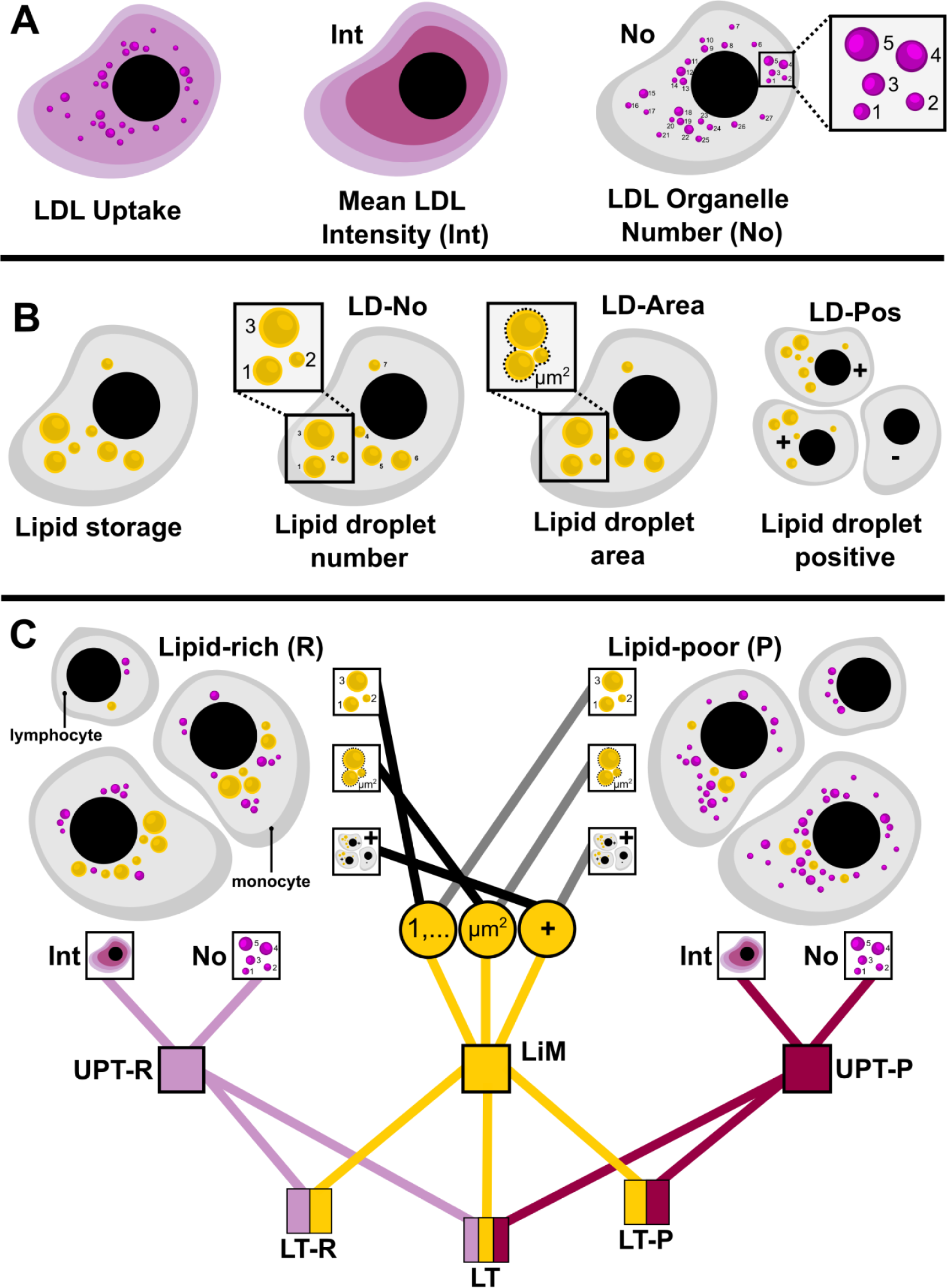
Multiparametric readouts for cellular lipid storage and trafficking. (A) Schematic presentation of fluorescent LDL uptake in cells and quantification as mean cell intensity (Int) and the number of LDL-filled organelles (No). (B) Quantification of cellular lipid storage by determining the number of lipid droplets (LD-No), the total cellular lipid droplet area (LD-Area) and the proportion of lipid droplet positive cells (LD-Pos). (C) Each of these readouts was measured in lipid-rich (R) condition (left) and lipid-poor (P) condition (right) in monocyte and lymphocyte populations. First, primary uptake readouts for lipid-rich and -poor conditions were combined into uptake scores (UPT-R and UPT-P) and lipid mobilization score was calculated by combining the ratios of lipid storage parameters in lipid rich to lipid poor conditions. Then uptake and mobilization scores were further combined into higher-order lipid trafficking (LT) scores (C).

In the lipid storage assay we analyzed lipid droplets in lipid-rich and -poor conditions, quantifying the number of lipid droplets (LD-No) and the total lipid droplet area (LD-Area) in every lymphocyte and monocyte as well as the percentage of lipid droplet-positive cells (LD-Pos) for lymphocyte and monocyte populations (Figure 1B). Both lymphocytes and monocytes tended to deplete the lipids stored in lipid droplets when challenged with lipid-poor conditions. We expressed this by dividing the values of lipid droplet parameters in lipid-rich and -poor conditions and further combining these three values into a single lipid mobilization score (Figure 1C). Moreover, we combined lipid mobilization and LDL uptake scores into composite lipid trafficking scores for lipid-rich (LT-R) and -poor (LT-P) conditions, and into a single lipid trafficking (LT) score with all readouts combined (Figure 1C). This enabled us to look at individual cellular parameters as well as their combinations when integrating the data with physiological and clinical outcomes.

### Subject group characteristics

We have started the analysis by selecting 200 recipients of lipid-lowering therapy and 200 matched controls from the FINRISK 2012 Study for whom PBMC samples were stored in the Biobank. Relevant cellular readouts and nuclear magnetic resonance (NMR) metabolomics data could be obtained for 135 control subjects and 133 recipients of statin monotherapy (Supplementary Figure 1). We divided the group of statin recipients into sub-groups with increasing statin intensity based on reported LDL-C reductions from baseline achieved on different doses of simvastatin, atorvastatin and rosuvastatin.^29–31^ Of the subjects with available cellular readouts 60 received a statin dose enabling at least a 35% reduction in LDL-C from baseline (‘>35%’ group), 39 participants received therapy with a potential to reduce LDL-c by at least 40% (‘>40%’ group) and 30 people with the potential to reduce LDL-C by more than 45% (‘>45%’, group). We are referring to both ‘>40%’ and ‘>45%’ groups as recipients of high-intensity statin (HIS) medication (Supplementary Figure 1).

The percentage of males in the formed groups was 44% to 55% (Table 1). We observed small differences in BMI for statin users of the ‘>45%’ group (29.7 vs 27.8 in controls) (Table 1). There was no significant difference in waist and hip circumference in between the statin groups and controls (Table 1). As expected, subjects receiving statins had lower total cholesterol (4.61 mmol/L, statin vs. 5.65, controls) and LDL-c levels (2.51 mmol/L, statin vs. 3.48, controls) (Table 1).

**Table 1.** Baseline characteristics of the analyzed subject groups.

|  | **CONTROL(-)** | **STATIN(+)** | **>35%** | **>40%** | **>45%** |
| --- | --- | --- | --- | --- | --- |
| Subject number | 135 | 133 | 60 | 39 | 30 |
| Sex, male, n (%) | 69 (51%) | 70 (53%) | 33 (55%) | 17 (44%) | 14 (47%) |
| **Mean (SEM):** |  |  |  |  |  |
| Age, years | 62 (0.7) | 63.2 (0.7) | 63 (0.9) | 64.2 (1.0) | 64 (1.1) |
| BMI | 27.8 (0.4) | 28.6 (0.4) | 28.5 (0.6) | 28.9 (0.8) | 29.7 (0.9)* |
| Waist circumference, cm | 96.7 (1.1) | 98.7 (1.1) | 98.4 (1.5) | 98.7 (1.9) | 100.8 (2.2) |
| Hip circumference, cm | 102.7 (0.8) | 103.4 (0.9) | 102.5(1.3) | 104 (1.7) | 105.9 (2.1) |
| LDL-C, mmol/L | 3.48 (0.07) | 2.51 (0.07)+ | 2.41 (0.1)+ | 2.51 (0.14)+ | 2.52 (0.16)+ |
| Total cholesterol, mmol/L | 5.65 (0.08) | 4.61 (0.08)+ | 4.52 (0.13)+ | 4.64 (0.18)+ | 4.64 (0.23)+ |
| Triglycerides, mmol/L | 1.5 (0.1) | 1.54 (0.08) | 1.6 (0.13) | 1.7 (0.17) | 1.74 (0.22) |

BMI, body mass index; LDL-C, low density lipoprotein cholesterol; Each of the statin groups are compared to the control group. \* $p<0.05$ , \*\* $p<0.01$ , + $p<0.001$ .

### Association of lipid trafficking scores with LDL-cholesterol, apolipoprotein(B) and total cholesterol

Next, we investigated whether cellular readouts on lipid trafficking are associated with circulating lipid markers such as LDL-C, total cholesterol and apolipoprotein(B) (APO-B) for the different subject groups. Similar to our previous observations from FH patients, monocyte LDL uptake parameters showed a negative correlation with plasma LDL-C for statin recipients (Figure 2A and Supplementary Figure 2A,B).^21^ This correlation was more pronounced for the quantification of LDL-filled organelles (LDL-No) (Figure 2A) as compared to cellular LDL intensity (LDL-Int) (Supplementary Figure 2A) when measured in lipid-poor condition. Stronger associations were also seen in measurements in lipid-poor conditions as compared to lipid-rich conditions (Figure 2A, Supplementary Figure 2B) and when quantified in monocytes (Figure 2A, Supplementary Figure 2A) as compared to lymphocytes (Supplementary Figure 2C). The correlation strength for both LDL-No and LDL-Int with LDL-C increased when shifting towards higher statin intensity, except for the ‘>45%’ group. Similar correlations were obtained for total cholesterol and APO-B (Figure 2C). For the ‘>40%’ statin group, LDL-No explained 16% of the variability of the circulating LDL-C (coefficient of determination R^2^=0.16, p=0.01), 14% of the total cholesterol (R^2^=0.14, p=0.02) and 13% of APO-B (R^2^=0.13, p=0.02).

**Figure 2.**
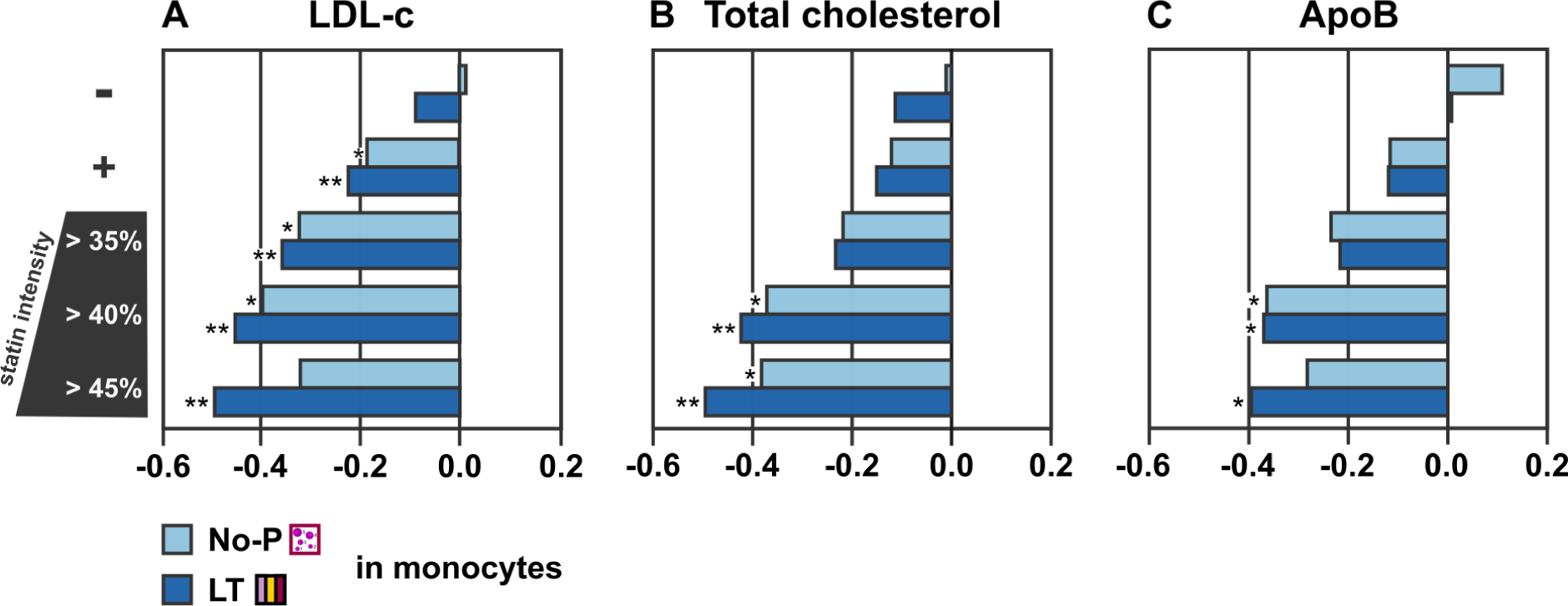
Cellular scores are associated with blood lipid markers for statin recipients. Both primary monocyte readouts such as LDL-filled organelle number in lipid-poor conditions (No-P) and combined lipid trafficking scores (LT) correlate negatively with circulating LDL cholesterol (A), total cholesterol (B) and Apolipoprotein(B) (C) in subjects on statin monotherapy. Minus sign represents control group (n=135) with no lipid-lowering medication use and plus sign represents subjects on statin monotherapy (n=133). The next three groups are for subjects receiving statin of moderate intensity and higher (>35%, n=60), subjects on high and very high intensity statin (>40%, n=39) and subjects on very high intensity statin therapy (>45%, n=30). * - p<0.05, ** - p<0.01.

The lipid mobilization score provided weak associations with LDL-C across the different groups and only saw an increase in the ‘>45%’ group (Supplementary Figure 3D). Combination of the lipid uptake scores with the lipid mobilization score into the lipid trafficking (LT) score (Figure 1C) provided stronger associations with LDL-C than LDL-No, with consistent increases for each statin intensity group (Figure 2A) and explaining 25% of the variability of LDL-C for the ‘>45%’ group (R^2^=0.25, p=0.005).

### Lipid trafficking scores associate with a pro-atherogenic lipoprotein profile

To investigate whether alterations of cellular lipid trafficking impacts also other lipid particles and species, we extended our study with NMR metabolomics, providing access to 213 biomarkers, including concentration and composition of 14 lipoprotein particle classes.

Based on our observations with LDL-C we utilized the LT score for correlations with the NMR metabolomics data. For control subjects, LT score showed a weak negative association with cholesterol ester and triglyceride concentration in large HDL particle fractions (Figure 3A) and with sphingomyelin (Figure 3C). When focusing on the statin group with an LDL-C-lowering potential of more than 40%, LT score correlated negatively with XS-VLDL, IDL, L-LDL, M-LDL, and S-LDL particle concentrations (Figure 3B). Higher cholesterol ester concentrations, in particles from S-VLDL to S-LDL were observed for subjects from the ‘>40%’ group with a low LT score, whilst associations with triglycerides were less pronounced (Figure 3B). We also observed correlations with the % lipid distribution across lipid particles. ‘>40%’-subjects with lower LT score displayed a higher share of cholesterol ester in small VLDL, IDL and LDL particles, whilst in large and medium sized HDL particles the share dropped (Figure 3B). Vice-versa, % triglycerides were lower in small VLDL and IDL particles for subjects with low LT score from the ‘>40%’ group (Figure 3B).

**Figure 3.**
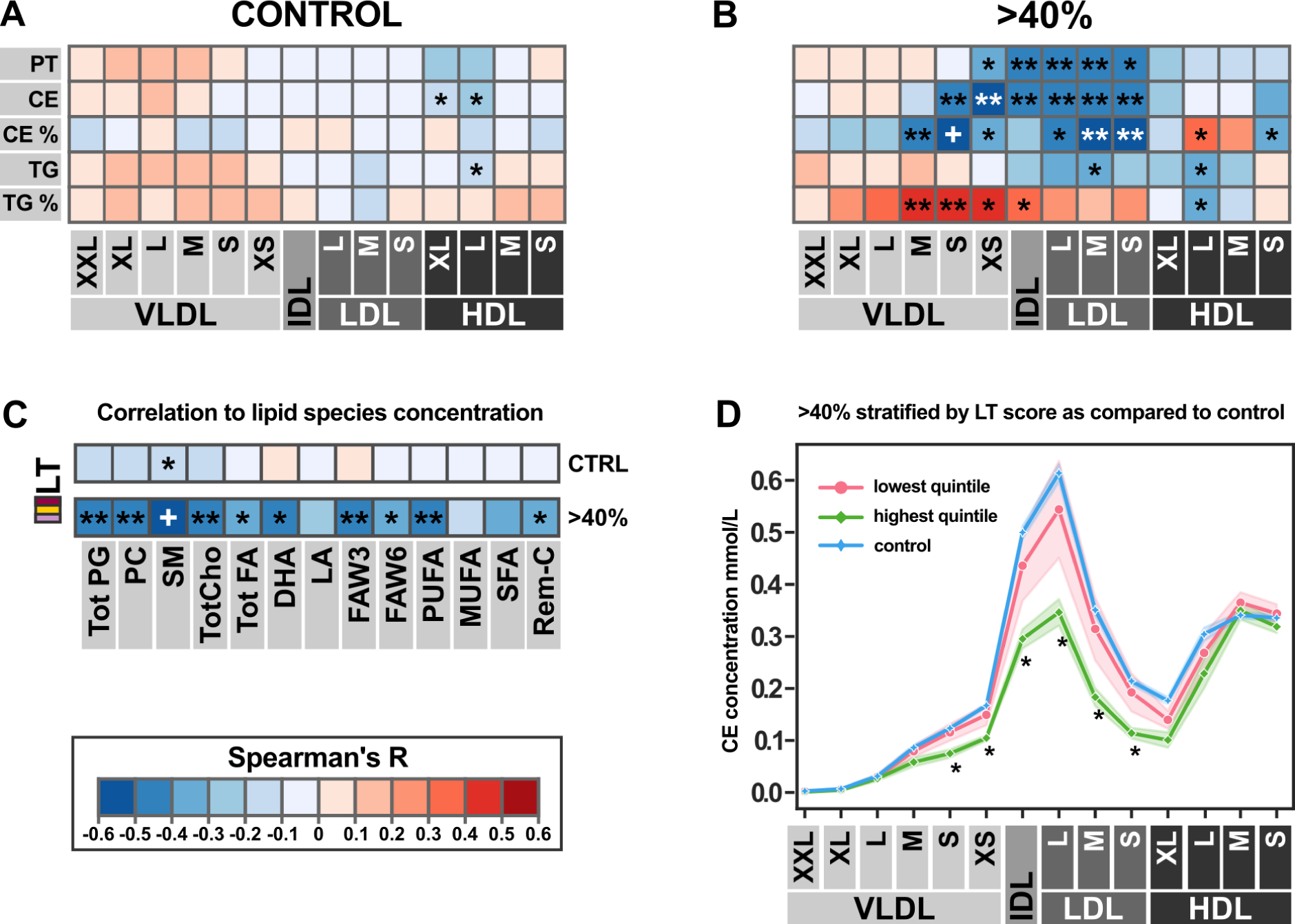
Low lipid trafficking scores are associated with a pro-atherogenic lipoprotein profile for high-intensity statin recipients. (A) Correlation heatmap of combined lipid trafficking (LT) score to different lipid concentrations in 14 lipoprotein subclasses in control subjects. PT stands for total particle concentration, CE – cholesterol ester concentration, CE % – percent of cholesterol ester in particles, TG – triglyceride concentration and TG % – percent of triglycerides in particles. Heatmap colors correspond to *r_s_*range and are explained at the bottom left corner of the figure. (B) Correlation heatmap of LT score to 14 lipoprotein subclasses in high-intensity statin recipients ‘>40%’ group. (C) Correlation heatmap of LT score to circulating concentration of different lipid species in control individuals versus high-intensity statin recipients ‘>40%’ group. The species are as follows: total phosphoglycerides (Tot PG), phosphatidylcholine and other cholines (PC), sphingomyelins (SM), total cholines (TotCho), total fatty acids (Tot FA), 22:06 docosahexaenoic acid (DHA), 18:02 linoleic acid (LA), omega-3 and omega-6 fatty acids (FAW3 and FAW6), polyunsaturated fatty acids (PUFA), monounsaturated fatty acids (MUFA), saturated fatty acids (SFA), remnant cholesterol (Rem-C). (D) The lowest quintile of LT scores for high-intensity statin users shows a cholesterol ester profile which is similar to controls, whilst subjects within the highest quintile have significantly reduced CE concentrations. * - p<0.05, ** - p<0.01, + - p<0.001

Besides cholesterol esters, LT score also negatively correlated with sphingomyelin concentration for the ‘>40%’ group (Figure 3C). Other lipid species were also increased for subjects with low LT, but association was less profound as compared to sphingomyelin.

Whilst ‘>40%’ subjects within the highest quintile of the LT score achieved substantial reductions in cholesterol ester concentrations of S-, XS-VLDL, IDL and LDL particles (Figure 3D), those within the lowest quintile did not and displayed cholesterol ester concentrations equivalent to subjects without cholesterol-lowering medication (Figure 3D).

### Lipid trafficking scores integrated with polygenic risk scores

Previously, we showed that cellular lipid trafficking scores did not associate with a polygenic risk score for LDL (LDL-PRS) for individuals without cholesterol-lowering medication.^21^ Here we confirm this observation with a larger set of individuals (Figure 4A). On the other hand, the combined LDL uptake score in lipid-poor conditions (UPT-P) correlated with LDL-PRS for subjects of the ‘>40%’ group, whilst the lipid mobilisation score and lipid trafficking score did not (Figure 4A). LDL-PRS correlated with LDL-C for statin users and, interestingly, the association strength increased within higher statin intensity groups (Figure 4B), except for the ‘>45%’ group, similar to the cellular LDL-No scores (Figure 2A). Based on these results we evaluated whether combination of LT score with LDL-PRS would lead to a better association with LDL-C as compared to the individual scores. The combined score showed significant associations with LDL-C for all statin groups and also control individuals (Figure 4B). For the ‘>45%’ group the combined score explained 29.7% of the LDL-C variability (R^2^=0.297, p=0.002), a 5.1% increase over the LT score (Figure 2A) and 20.6% over the LDL-PRS (R^2^=0.09, p=0.1). Then, we extended this analysis to the NMR metabolomic data (Figure 4C). Similar to the lipid trafficking score, a poor LDL-PRS associated with pro-atherogenic lipoprotein particles, with higher cholesterol ester and triglyceride content for subjects of the ‘>40%’ group (Figure 4C). Even though the % distribution of cholesterol esters and triglycerides was more affected by the LT score (Figure 4C, Figure 3B) combination of the LDL-PRS with the lipid trafficking score further improved the association strength with the pro-atherogenic lipoprotein profile (Figure 4D), explaining, for example, up to 37.7% of the variability (R^2^=0.377, p<0.001) of cholesterol ester concentration in XS-VLDL particles of ‘>40%’ subjects.

**Figure 4.**
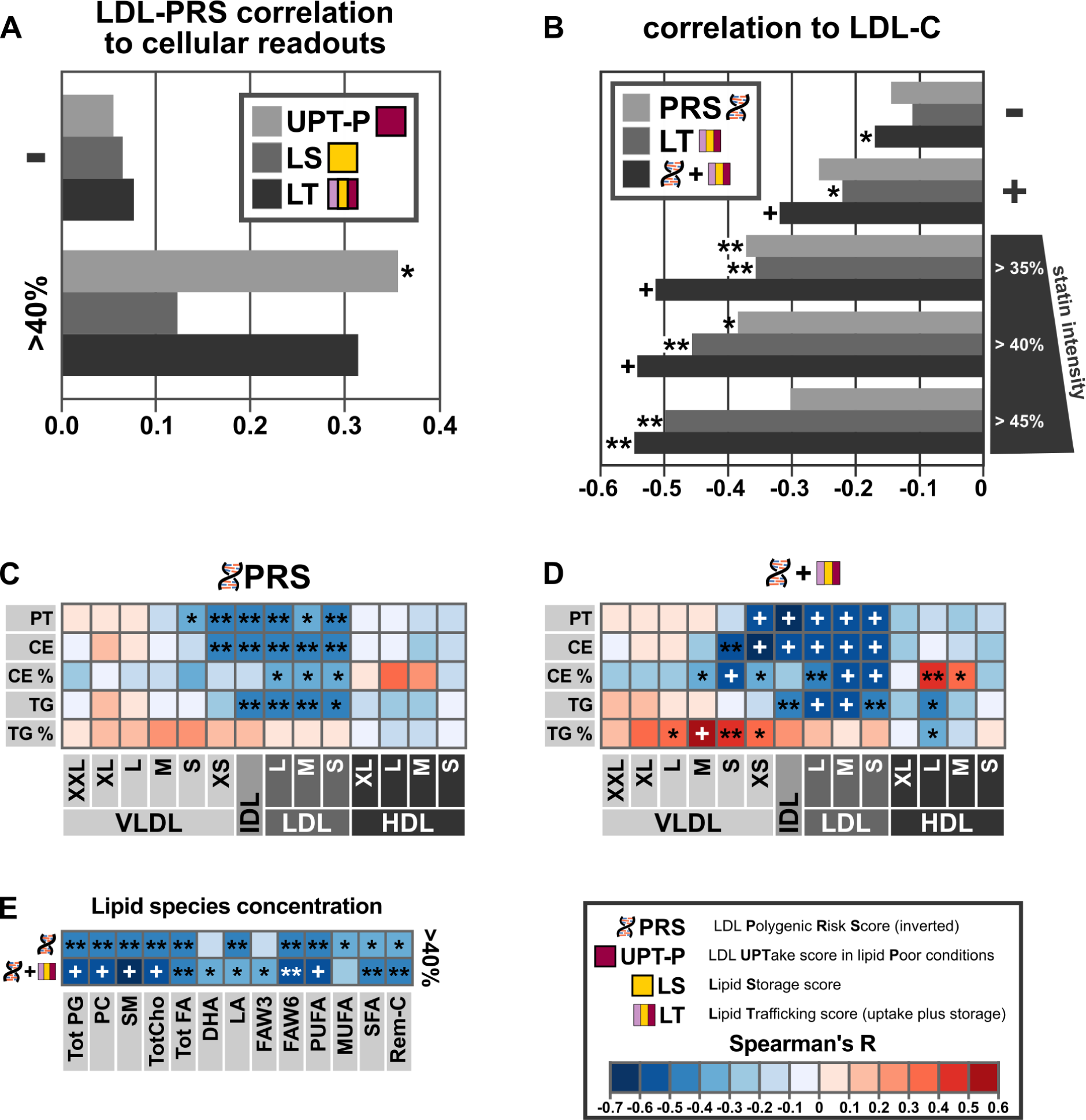
Cellular scores can be combined with LDL-PRS to improve the associations with LDL-c and other lipids. (A) Cellular LDL uptake (UPT-P) correlates with LDL-PRS (inverted) in high-intensity statin users (>40%, n=39) but not in control subjects (CTRL, n=134). (B) Combining LDL-PRS (inverted) and LT score into one combined score improves associations to LDL-c concentration in control subjects (-, n=134), statin users (+, n= 131), and subject groups with increasing statin intensity (>35%, n=59), (>40%, n=39) and (>45%, n=30). (C) Correlation heatmap of LDL-PRS (inverted) score to 14 lipoprotein subclasses in high-intensity statin recipients ‘>40%’ group. PT stands for total particle concentration, CE – cholesteryl ester concentration (CE), CE % – percent of CE in particles, TG – triglyceride concentration and TG % - percent of TG in particles. Heatmap colors correspond to Spearman’s rank range and are explained at the bottom right corner of the figure. (D) Correlation heatmap of combined score of LDL-PRS with LT to 14 lipoprotein subclasses in high-intensity statin recipients ‘>40%’ group. (E) Correlation heatmap of LDL-PRS (inverted) score and combined LDL-PRS-LT score to circulating concentration of different lipid species in high-intensity statin recipients ‘>40%’ group. The species are as follows: total phosphoglycerides (Tot PG), phosphatidylcholine and other cholines (PC), sphingomyelins (SM), total cholines (TotCho), total fatty acids (Tot FA), 22:06 docosahexaenoic acid (DHA), 18:02 linoleic acid (LA), omega-3 and omega-6 fatty acids (FAW3 and FAW6), polyunsaturated fatty acids (PUFA), monounsaturated fatty acids (MUFA), saturated fatty acids (SFA), remnant cholesterol (Rem-C). * - p<0.05, ** - p<0.01, + - p<0.001.

### Lipid trafficking scores enable stratification for LDL target level attainment

Next, we investigated whether lipid trafficking scores and LDL-PRS could enable the definition of subject sub-groups who are at higher or lower odds to achieve their LDL-C treatment goals. According to the 2011 ESC/EAS guidelines^32^ that were in use during the sample collection, LDL-C goal of <1.8 mmol/l was recommended for very high cardiovascular risk and a goal of <2.5 mmol/l for high cardiovascular risk. We adopted these recommendations for our goal-attainment analysis defining subjects with a history of myocardial infarction or stroke as being at very high risk and subjects without such a history at high risk. Subjects in each group were then divided into quintiles for the different LT scores and we evaluated whether subjects within the highest or lowest quintile achieved LDL-C target levels. Whilst all LT scores showed the same tendency, LT-R (a combination of LDL uptake scores quantified from monocytes in lipid-rich conditions with the lipid mobilization score) was the best at stratifying subjects for reaching their LDL target levels. We then looked at the performance of the LT-R, LDL-PRS and a combined LDL-PRS/LT-R score (Figure 5).

**Figure 5.**
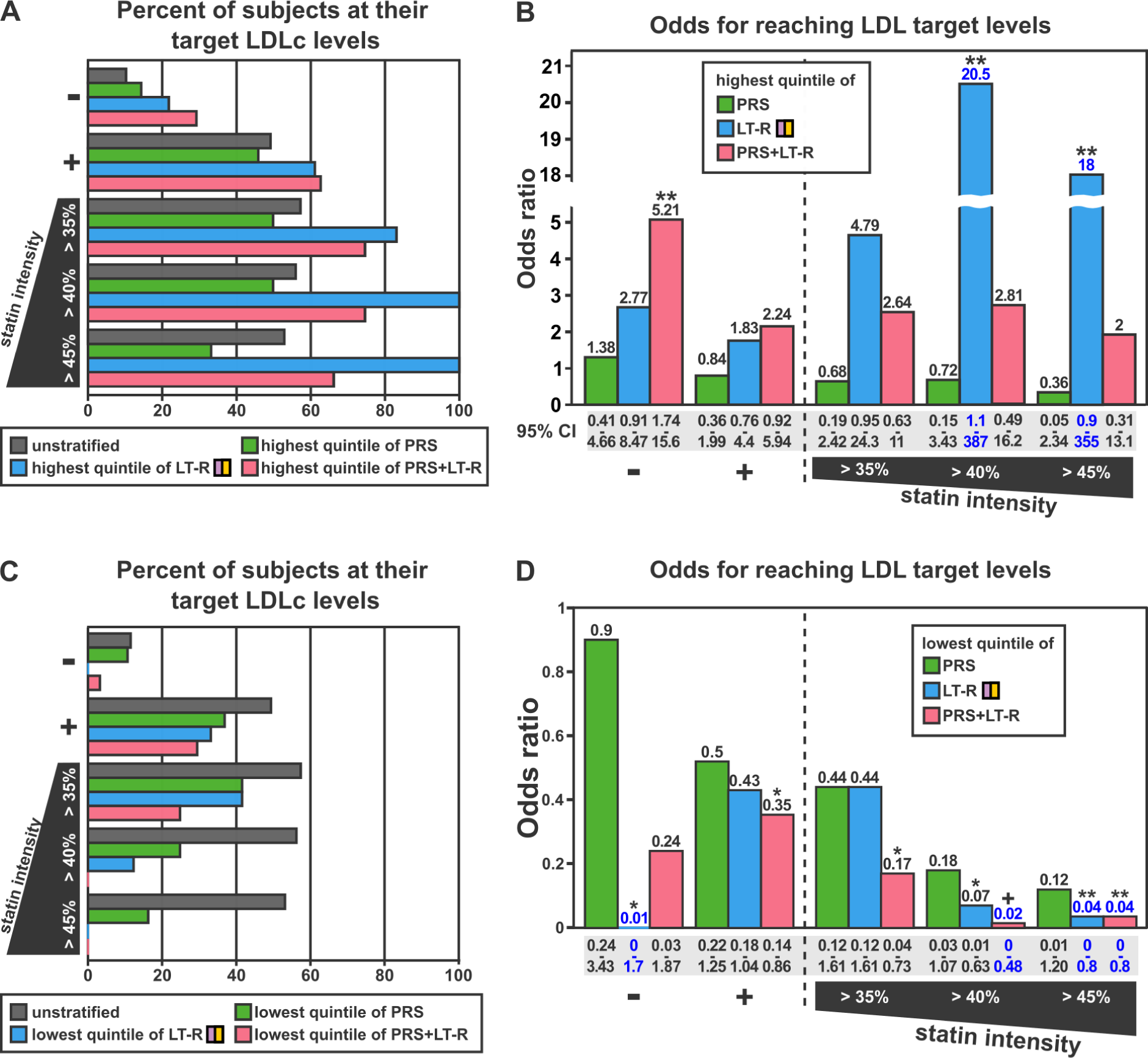
Using cellular scores to stratify subjects for LDL goal attainment. (A) Percent of subjects achieving their LDL-C goals when being within the highest quintile of the LDL-PRS (green), lipid trafficking score (LT-R) (blue) or combined LDL-PRS/LT-R score (magenta) as compared to subjects without stratification (gray). (B) Odds ratios for achieving LDL-C target levels for subjects within the highest quintile of the LDL-PRS (green), LT-R(blue) and combined LDL-PRS/LT-R score for the same subject groups as in (A). (C) Percent of subjects achieving their target goals when being within the lowest quintile of the LDL-PRS (green), lipid trafficking score (LT-R) (blue) or combined LDL-PRS/LT-R score (magenta) as compared to subjects without stratification (gray). (D) Odds ratios for achieving LDL-C target levels for subjects within the lowest quintile of the LDL-PRS (green), LT-R (blue) and combined LDL-PRS/LT-R score for the same subject groups as in (C). Control (-) group had 134 subjects and 27 in focus quintiles, statin group (-) had 131 subjects and 27 in focus quintiles, ‘>35%’ group had 59 subjects and 12 in focus quintiles, ‘>40%’ group consisted of 39 subjects and 8 per analyzed quintile, ‘>45%’ group has a total of 30 subjects with 6 in each quintile. Blue text in (B) and (C) indicates estimated odds ratios and confidence intervals. * - p<0.05, ** - p<0.01, + - p<0.001.

Those subjects within the highest LT-R quintile displayed better goal attainment than unstratified subjects across the different study groups and goal attainment further increased in high-intensity statin groups (Figure 5A). For both ‘>40%’ and ‘>45%’ groups all subjects within the highest LT-R quintile achieved their LDL target goals, therefore we used estimates of the odds ratios for these subgroups (Figure 5 A,B). Looking at those subjects within the highest LDL-PRS quintile only provided marginal improvements for goal attainment above unstratified subjects of the control group (Figure 5A,B). Also, combining LT-R and LDL-PRS did not provide added value for subjects on statin medication. However, for those without statin medication, combining LT-R and LDL-PRS enabled the definition of a subject subgroup with a five-fold higher odds to achieve target LDL-C levels (OR=5.2, 95% CI [1.74, 15.6]) compared to the rest of the group (Figure 5B).

Without statin medication, none of the subjects within the lowest LT-R quintile achieved LDL-C target levels (Figure 5C, D). Goal attainment increased for subjects of the same LT-R quintile when looking at the entire statin group, but then dropped for subjects in groups with higher statin intensity. In the ‘>40%’ group, only 12.5% of subjects within the lowest LT-R quintile achieved their LDL-C goal and no subjects within the lowest quintile achieved their goal in the ‘>45%’ group (estimated OR=0.02, 95% CI [0.001, 0.475] P<0.001). Also individuals within the lowest LDL-PRS quintile were at lower odds to achieve their LDL-C targets (Figure 5C,D), but LT-R showed stronger effects. Combination of LDL-PRS and LT-R improved the definition of subjects who did not achieve LDL-C target levels, especially in the ‘>35%’ and ‘>40%’ statin groups.

### Lipid trafficking scores and cardiovascular disease

During a seven-year follow-up of the 265 subjects, eleven (four in controls and seven in statin group) were diagnosed with myocardial infarction (MI) and ten with stroke (five in each of the two groups), resulting in 21 cardiovascular events. This allowed us to get a first impression whether cellular scores could aid in cardiovascular risk assessment.

Both control subjects and statin recipients within the lowest quintile of lipid mobilization score were at higher odds to experience a cardiovascular event as compared to the rest of their group with increasing effect size for higher statin intensity (Figure 6). There was a significant difference in the odds to experience a CV event between the lowest quintile of lipid mobilization score and the rest of the subjects in both the ‘>40%’ group and the ‘>45%’ group (Figure 6). Combination of the lipid mobilization score with the LDL uptake score from lipid-poor conditions increased the odds ratio of experiencing a cardiovascular event to 30 (95%CI [2.65, 339.75], p=0.004) for subjects in the lowest quintile of the ‘>40%’ group, as compared to the rest of the group.

**Figure 6.**
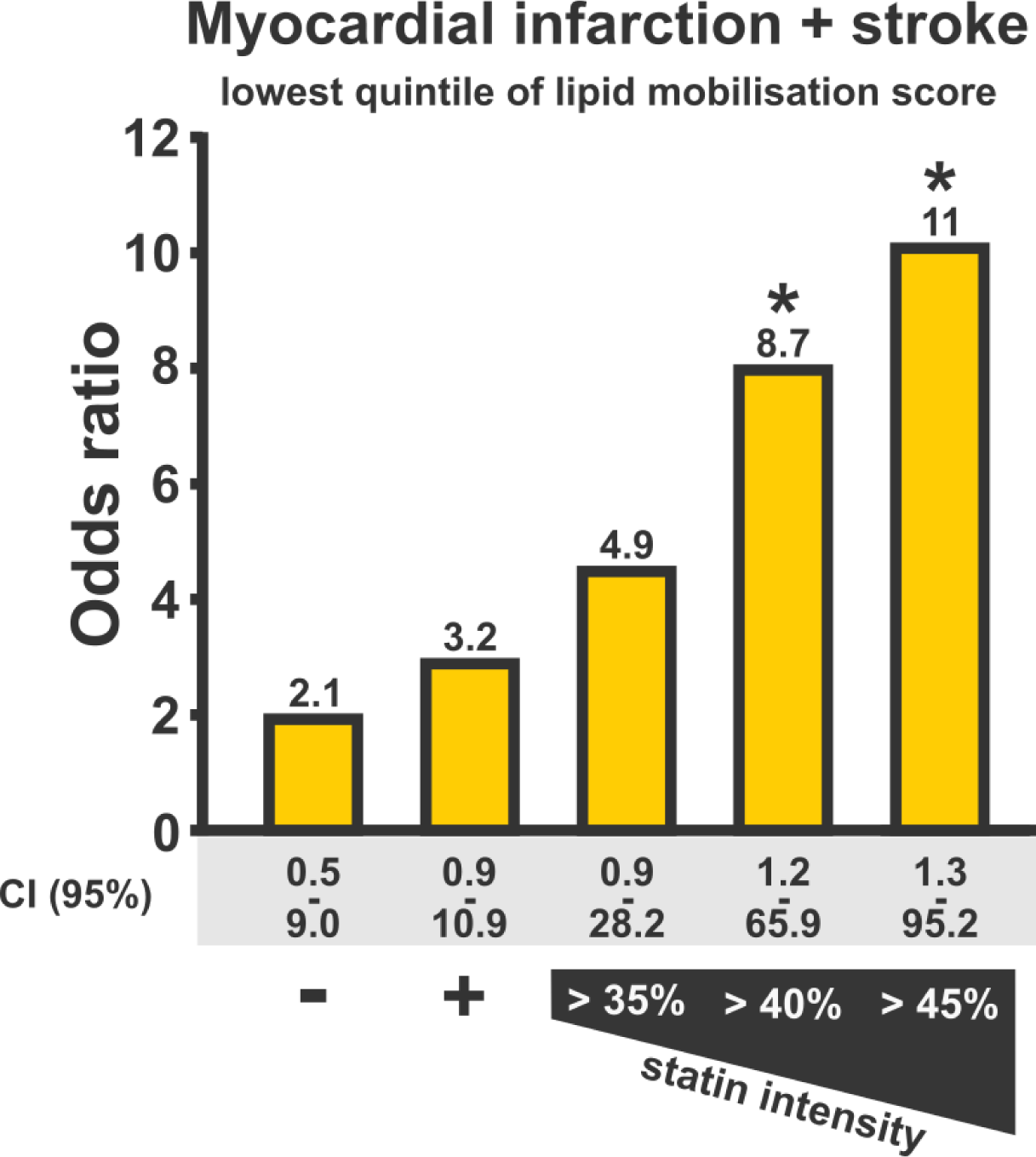
Lipid mobilization score in risk assessment for myocardial infarction and stroke. Subjects in the lowest quintile of lipid mobilization score have increased odds of experiencing a cardiovascular event as compared to the rest of the group. The odds ratio is higher with a concomitant increase in statin intensity. Control (-) group had 134 subjects (27 in focus quintile), statin group (-) had 131 subjects (27 in focus quintile), ‘>35%’ group had 59 subjects (12 in focus quintile), ‘>40%’ group consisted of 39 subjects (8 in focus quintile) and ‘’>45%’ group had a total of 30 subjects(6 in each quintile). * p<0.05.

## Discussion

We performed a systematic functional cellular analysis for a set of subjects from a representative, cross-sectional population survey, observing considerable interindividual variability and widespread defects in cellular LDL uptake and lipid mobilization in leukocyte subpopulations. We further combined individual LDL-uptake and lipid mobilization readouts into one lipid trafficking (LT) score. For subjects receiving high-intensity statin (HIS) medication, lower cellular LDL uptake and LT score were associated with higher circulating concentrations of LDL-C, IDL, small VLDL particles and cholesterol ester content across APO-B containing lipid particle subclasses. Elevated LDL-C is responsible for the formation of atherosclerotic plaques and progression of cardiovascular disease.^1^ Besides LDL, small VLDL and IDL can cross the endothelial cell barrier and remnant cholesterol is a known driver of atherosclerosis.^33,34^ We further observed an inverse association between lipid trafficking score and sphingomyelin concentration in HIS recipients. A higher amount of sphingomyelin in LDL particles renders them more prone to aggregation and stimulates atherosclerosis.^35^ It appears that subjects with poor lipid trafficking scores have limited capacity to benefit from statin monotherapy, which is also reflected in them being at higher odds for experiencing an adverse cardiovascular event when compared to others on equivalent medication. Statin acts by blocking cholesterol synthesis and elevating the expression of liver LDL receptors.^16,28^ Subjects with reduced cellular capacity to uptake LDL and to utilize stored lipids may have a limited ability to upregulate LDLR and consequently efficiently clear pro-atherogenic lipoproteins upon statin therapy. This reasoning can be also supported by our observation that the association of LT score with LDL-C is strengthening with increase in statin intensity, reflecting greater reliance of subjects on cellular LDL uptake pathways in such conditions.

Similar to the lipid trafficking score, association strength of LDL-PRS with LDL-C increased with higher statin intensity. A potential explanation for this could be that a portion of the single nucleotide polymorphisms contained in the LDL-PRS is linked to cellular LDL uptake. Our observation that LDL uptake correlates with the LDL-PRS for HIS recipients supports this hypothesis. Apparently, LDL-PRS and lipid trafficking score have non-overlapping features as well, which would explain why a combination of the LDL-PRS with the lipid trafficking score provided a better association with pro-atherogenic lipoprotein particles and in some instances improved subject stratification for LDL-C goal attainment.

Our observations are supported by genetic studies which link loss-of-function variants of the LDL receptor to a higher incidence of cardiovascular disease.^36,37^ Whilst the detection of an LDLR loss-of-function variant implies defective LDL uptake, we directly quantify this cellular process with high-content microscopy of leukocyte subpopulations.

It is known that individuals from the general population can have a cellular LDL uptake similar to heterozygous familial hypercholesterolemia patients with loss-of-function LDLR variants.^27^ Our study supports these findings, and highlights for the first time the physiological and clinical consequences for subjects with lipid trafficking defects in the general population.

Interindividual variability in treatment outcomes for statin monotherapy was observed before^3,13,14^, also when statin adherence was monitored closely^15^. Consequently, it seems unlikely that the interindividual variability only stems from non-adherence. Several studies highlight difficulties in LDL-C goal attainment for patients on statin medication and in making the switch to combination therapy.^2,5^ Systematic quantification of cellular lipid trafficking pathways may aid in the identification of patients who are refractory to statin medication, remain at increased cardiovascular risk and are in need of combination therapy. This is supported by our observation that individuals with a poor lipid trafficking score do not achieve their LDL-C goals on HIS medication. On the other hand, all subjects with a good lipid trafficking score achieved their LDL-C goals on HIS therapy, a piece of information which may be important for physicians as well, providing additional justification for treatment selection and potentially improved acceptance by the patient.

It is important to keep in mind, however, that our current observations for HIS recipients are derived from a limited number of subjects from the Finnish population This might affect the applicability of these results to other populations, especially those with different ethnic backgrounds. The medication status of the subjects in this study was determined by their drug purchase patterns. Whilst this method gives a more detailed picture compared to the yes/no type of questionnaire initially available, it can only provide an approximation of real adherence.

Here we acquired a comprehensive set of functional cell data for 268 individuals from the FINRISK 2012 Study. The analysis strategy is based on a semi-automated workflow with fully automated microscopy and image analysis. This reduces researcher bias and increases the reproducibility of the results simultaneously making it possible to scale the analysis to higher throughput, enabling clinical applications in the future. This study highlights that intraindividual variation of biological processes should receive more attention in the treatment of hypercholesterolemia.

## Online Methods

### Study design and group definitions

We selected 200 recipients of lipid-lowering therapy and 200 matched controls from the FINRISK 2012 population survey for whom frozen PBMC samples were stored in THL Biobank. The case and control groups were selected based on the questionnaire answer for undergoing lipid-lowering therapy. Each group was chosen to include subjects with a broad range of LDL-c values and equal sex distribution; medication for blood pressure or diabetes was avoided when possible. The case-control matching was then done manually based on age, sex, bmi, after filtering out the pools for case population and control population. Direct matching to each case was performed.

We obtained laboratory lipid values (Total cholesterol, LDL-C, triglycerides, APO-B, APO-A1), age, sex, BMI, wais and hip circumference for all of the selected individuals. NMR metabolomics values were available for 398 subjects and polygenic risk scores could be calculated for 394 participants. For optimal performance of the cellular assays 1.6 million viable cells were required per each subject. We could obtain only one tube of cryopreserved PBMCs stored in the Biobank for each subject. The amount of viable cells per subject ranged from zero to 5.5 million, therefore we could obtain reliable cellular readout results from 273 samples (135 controls and 138 cases).

For a precise definition of the used lipid-lowering medication we utilized drug purchase history for 2002-2019 obtained from the Finnish Social and Health Data Permit Authority Findata. We selected subjects who purchased lipid-lowering medication during 6 months prior to sampling. The last two purchases before sampling defined the medication type. This strategy resulted in selection of 202 subjects of whom 191 were on statin monotherapy (simvastatin n=127, atorvastatin n=30, rosuvastatin n=22, fluvastatin n=3, lovastatin n=3, pravastatin n=6), 9 were on combination therapy or took other lipid-lowering medication (ezetimibe=2, fenofibrate=1, statin/ezetimibe=5, statin/ezetimibe/fenofibrate=1) and two subjects have switched statin type during the target time period. Since 94% of subjects were on either simvastatin, rosuvastatin or atorvastatin monotherapy we focused only on these three statin types in the further subgroup formation.

We further extracted the purchased statin doses from the records and defined statin intensity groups based on previously reported LDL-c reductions from baseline achieved with different doses of simvastatin, rosuvastatin and atorvastatin: >35% LDL-c reduction from the baseline (rosuvastatin 5-40 mg, atorvastatin 10-80 mg, simvastatin 40-80 mg), >40% (rosuvastatin 5-40 mg, atorvastatin 10-80 mg) and >45% (rosuvastatin 10-40mg, atorvastatin 20-80 mg).^29–31^ Therapy adherence was estimated by dividing the number of tablets in the package bought before sampling by the number of days elapsed until the next purchase. The average adherence was as follows (mean±SEM): statin group 0.86±0.03, ‘ >35%’ group 0.89±0.04, ‘>40%’ group 0.83±0.04 and ‘>45%’ group 0.83±0.05. 55-62% of subjects in each analyzed group reported having a history of taking blood pressure medications, additionally 4% of controls and 23-26% of statin recipients reported using diabetes medication (Supplementary Table 1).

We have conducted the initial association analyses for the subjects for whom both cellular data and NMR metabolomics values were both available. Before integration of cellular score with LDL-PRS, subjects missing the latter were also removed (Supplementary Figure 1).

### Definition of adverse cardiovascular events

Cardiovascular disease events were defined as myocardial infarction or ischemic stroke specified by I2X and I6X diagnosis codes from the International Statistical Classification of Diseases and Related Health Problems 10th Revision (Supplementary Methods). Both prevalent and incident CVD events were included and were extracted from hospital admission and primary care electronic health records.

### Human subject samples

Peripheral blood mononuclear cell (PBMC) samples were cryopreserved in 2012 as a part of Finnish population survey, FINRISK 2012, with written consent and ethical approval of the Hospital District of Helsinki and Uusimaa (permit 162/13/03/00/2011).^38^ The FINRISK 2012 sample collection was then transferred to THL Biobank in 2015. Frozen PBMC samples, pseudonymised donor-linked lipid values and survey data were obtained then from THL Biobank (www.thl.fi/biobank) and used under Biobank agreement (THLBB2020_7). A mixture of PBMCs from four healthy anonymous donors were used as standards for assay measurements. These cells were isolated by density-gradient centrifugation from buffy coat samples obtained through the Finnish Red Cross Blood Service (56/2019).

### PBMC recovery

Cryopreserved samples were thawed and cell viability was determined using trypan blue exclusion technique. Cells were then seeded into 96-well plates for 24-hour incubation with lipid-rich (R) medium (RPMI-1640 supplemented with 2mM L-Glutamine, 100 units/mL potassium penicillin, 100ug/mL streptomycin sulfate, 5mM HEPES and 1mM sodium pyruvate) containing 10% fetal bovine serum or lipid-poor (P) medium containing 5% lipoprotein-deprived serum. The cellular assays were performed in 384-well plate format on the next day.

### LDL uptake and lipid droplet assays

Fluorescent labeling of low-density lipoprotein particles with DiI^39–41^, LDL uptake and lipid droplet assays^21^ were described previously. DiI-LDL was spiked into the growth media to achieve a final concentration of 30 μg/mL and the cells were further incubated for 1 hour at 37°C. Subsequently, the cells were washed and resuspended with serum-free growth medium. Next, using an open-source robotic platform (Opentrons, New York, USA) the cells were transferred and adhered to coated 384-well imaging plates. Afterwards, cells were fixed with 4% PFA, washed and stained with nuclear marker DAPI (5 μg/ml) and cytoplasm marker CellMask™ DeepRed (0.5 μg/ml). For lipid droplet analysis the cells were fixed and stained in the same manner as in the LDL uptake assay except that the staining solution contained 5 μg/ml DAPI and 1μg/ml LD540 (Princeton BioMolecular Research).^42^ Images were acquired for both assays with a PerkinElmer OperaPhenix automated spinning disc confocal microscope using a 40x or 63x water immersion objective. Each assay was repeated twice and two PBMC standard samples were included in every experiment. Each analysis batch included matched cases and controls.

### Image processing and quantification

Raw confocal 3D image stacks were automatically deconvolved and collapsed into the maximum intensity projections using a custom-made Python tool (https://github.com/lopaavol/OPutils). Then cell nuclei, cytoplasm and organelles were segmented automatically using CellProfiler.^43^ Several images from each plate were sampled and inspected by the experimenter for potential segmentation errors to ensure segmentation quality and to filter out imaging artifacts.

### Genotyping, imputation and LDL-C polygenic risk score (LDL-PRS)

We obtained genotypes for 394 of our 400 study individuals from THL Biobank for which single nucleotide polymorphism (SNP) array and centrally imputed genotype data were available. The FINRISK 2012 samples were genotyped using the Illumina HumanCoreExome-24 v1.1 and Affymetrix Axiom FinnGen1.r1 and FinnGen1.r2 arrays. The genotype imputation workflow with pre- and post-imputation quality control steps was carried out as described in the following protocol: dx.doi.org/10.17504/protocols.io.xbgfijw.

We used PLINK 2.0^44^ (www.cog-genomics.org/plink/2.0/) to first convert PED and MAP files obtained from THL Biobank to BED, BIM and FAM files, and then calculated a genome-wide LDL-PRS using weights for a LDL-PRS calculated in FinnGen^45^ study Data Freeze 7 (available at: https://www.pgscatalog.org/score/PGS002764/). Our final LDL-PRS included 1,086,403 variants and could be calculated to 394 out of 400 subjects. Next, we normalized LDL-PRSs to zero to one range. Contrary to cellular scores, LDL-PRS was positively associated with LDL-c levels in our study (Supplementary Figure 3) and in previously reported FINIRSK 2012 cohort analysis.^46^ Therefore, to allow for comparability of LDL-PRS to cellular lipid trafficking scores and their further integration we inverted LDL-PRS for all the downstream analyses.

### Subject health data retrieval

General information about subjects’ age, sex, weight, BMI, waist and hip circumference, lifestyle and medication status (lipid-lowering, diabetes and blood pressure) as well as standard laboratory lipid values and NMR metabolomics values were retrieved in conjunction with samples from THL Biobank. Additionally the pseudonymised medical records related to cardiometabolic disorders for each subject were obtained for years 2002– 2019 from Finnish primary care register (Avohilmo) and national social and healthcare data collection and reporting system (Hilmo). Records of purchase and reimbursement of lipid-lowering medications for years 2002–2019 were obtained from the Social Insurance Institution of Finland (Kela). All data extraction procedures were done under permit (THL/4640/14.02.00/2020) and handled by Findata.

### Data processing and statistical analysis

Data was processed and visualized using Python standard libraries (Python Software Foundation, www.python.org) and the following packages: Pandas,^47^ Numpy,^48^ Scipy,^49^ Matplotlib,^50^ Scikit-Learn,^51^ Seaborn^52^. Single-cell readouts were filtered for cell size and mean intensity outliers with 5 SD from mean being the cut-off point. On average several hundred monocytes and several thousand lymphocytes were quantified in each well. Wells with less than 50 cells were discarded from the analysis. We further obtained mean per-well values for intensity and organelle numbers. For each of lipid-rich (R) and lipid-poor (P) conditions we analyzed four wells in two independent experiments. Absolute values were then normalized to the included control samples. LDL uptake and lipid mobilization for the entire dataset were additionally rescaled to zero to one range before combining into lipid trafficking scores.

Statistical analyses were performed with python package scipy.stats. For pairwise comparisons of group means reported in Table 1 the Student’s T-test was used. To determine the correlations between cellular scores and lipid values the Spearman rank-order correlation coefficients and corresponding asymptotic P values were calculated. To test the difference between lipid species concentrations in controls versus subgroups of statin recipients (Figure 3D) we used Mann-Whitney U test. Odds ratios were calculated using 2×2 contingency tables and corresponding P values were calculated using Fisher’s exact test. The 95% confidence intervals (CI) were calculated using the standard formula for CI for odds ratios. Due to small sample size in some cases the contingency tables contained zero values and therefore produced zero or infinite odds ratios, to calculate an estimated odds ratio in these cases a value of 0.5 was added to each value in a 2×2 table.

## Acknowledgements

The samples and sample-related data used for the research were obtained from THL Biobank (study number: THLBB2020_7). We thank all study participants for their generous participation at THL Biobank and FINRISK 2012 Study.

## Sources of Funding

Grants from the Academy of Finland 328861 and 325040, Business Finland (Research to Business) 1821/31/2021, Magnus Ehrnrooth and the Foundation for Cardiovascular Research to S.G.P. Grants from Academy of Finland Center of Excellence in Complex Disease Genetics 312062, Academy of Finland 285380, the Finnish Foundation for Cardiovascular Research and the Sigrid Jusélius Foundation to S.R. Funding from the Doctoral Programme in Population Health, University of Helsinki to M.T.

## Competing interests statement

A patent application covering the use of the here-suggested stratification methods has been filed (application number: FI2021050861W·2021-12-10), in which University of Helsinki is the applicant and Simon G. Pfisterer and Elina Ikonen are inventors. The application is currently at the stage of individual filing in selected countries. The technology for the cellular analysis covered in patent application is used in this manuscript and has also been previously published as a proof-of-concept study for familial hypercholesterolemia patients.

## Author contribution

S.G.P. and I.H. designed the study. I.H. and M.M.I. performed experiments and analyzed the cellular data. M.T. and S.R. performed the calculations of polygenic risk scores. S.G.P and I.H. interpreted the results. S.G.P. and I.H. wrote and revised the manuscript. S.R and M.T. have critically reviewed the manuscript. All authors have approved the manuscript for publication.

## Data availability statement

The data used in this research includes sensitive personal health information and is therefore under restricted access. Biological samples and linked laboratory lipid values, questionnaire answers and biometrics as well as genetic data underlying this article were provided by THL Biobank (www.thl.fi/biobank) and used under Biobank agreement THLBB2020_7. Access to the data can be obtained from the THL Biobank through the standard application procedure. The register-based data including diagnoses, causes of death and medication purchase history used in this study can be obtained from the Finnish Social and Health Data Permit Authority Findata by submitting a data permit application. Description of the register data used in the study (names of registers and attribute codes) can be obtained from the corresponding author.

## Notes

### Competing Interest Statement

A patent application covering the use of the here-suggested patient stratification methods has been filed (application: WO2022123120A1), in which University of Helsinki is the applicant and S.G.P. is an inventor.

### Author Declarations

Peripheral blood mononuclear cell (PBMC) samples were cryopreserved in 2012 as a part of Finnish population survey, FINRISK 2012, with written consent and ethical approval of the Hospital District of Helsinki and Uusimaa (permit 162/13/03/00/2011). The FINRISK 2012 sample collection was then transferred to THL Biobank in 2015. Frozen PBMC samples, pseudonymised donor-linked lipid values and survey data were obtained then from THL Biobank (www.thl.fi/biobank) and used under Biobank agreement (THLBB2020_7). The rigister-based pseudynymised data was obtained under permit (THL/4640/14.02.00/2020) from the Finnish Social and Health Data Permit Authority Findata.

### Summary of Updates

Abstract was shortened. Discussion was rewritten to add clarity.

## References

1. Borén, J. et al. Low-density lipoproteins cause atherosclerotic cardiovascular disease: pathophysiological, genetic, and therapeutic insights: a consensus statement from the European Atherosclerosis Society Consensus Panel. Eur. Heart J. 41, 2313–2330 (2020).

2. Ray, K. K. et al. EU-Wide Cross-Sectional Observational Study of Lipid-Modifying Therapy Use in Secondary and Primary Care: the DA VINCI study. Eur. J. Prev. Cardiol. 28, 1279–1289 (2021).

3. Ridker, P. M., Mora, S., Rose, L., & JUPITER Trial Study Group. Percent reduction in LDL cholesterol following high-intensity statin therapy: potential implications for guidelines and for the prescription of emerging lipid-lowering agents. Eur. Heart J. 37, 1373–1379 (2016).

4. Ballantyne, C. M. Achieving greater reductions in cardiovascular risk: lessons from statin therapy on risk measures and risk reduction. Am. Heart J. 148, S3–S8 (2004).

5. Ray, K. K. et al. Treatment gaps in the implementation of LDL cholesterol control among high- and very high-risk patients in Europe between 2020 and 2021: the multinational observational SANTORINI study. Lancet Reg. Health Eur. 29, 100624 (2023).

6. Makhmudova, U., Wolf, M., Willfeld, K., Beier, L. & Weingärtner, O. Different Perspectives of Patients and Physicians on LDL-C Target Achievement in the Treatment of Hypercholesterolemia: Results on Secondary Prevention from the German PROCYON Survey. Adv. Ther. 40, 460–473 (2023).

7. Vandewalle, L., Duchi, F., Verhelle, K. & Vanacker, P. Suboptimal lipid management in patients with acute ischemic stroke. Clin. Neurol. Neurosurg. 229, 107717 (2023).

8. Mitani, H. et al. Achievement Rates for Low-Density Lipoprotein Cholesterol Goals in Patients at High Risk of Atherosclerotic Cardiovascular Disease in a Real-World Setting in Japan. J. Atheroscler. Thromb. advpub, 63940 (2023).

9. Katzmann, J. L. et al. Trends in Ezetimibe Prescriptions as Monotherapy or Fixed-Dose Combination in Germany 2012–2021. Front. Cardiovasc. Med. 9, 912785 (2022).

10. Cohen, J. D., Cziraky, M. J., Jacobson, T. A., Maki, K. C. & Karalis, D. G. Barriers to PCSK9 inhibitor prescriptions for patients with high cardiovascular risk: Results of a healthcare provider survey conducted by the National Lipid Association. J. Clin. Lipidol. 11, 891–900 (2017).

11. Hirsh, B. J., Smilowitz, N. R., Rosenson, R. S., Fuster, V. & Sperling, L. S. Utilization of and Adherence to Guideline-Recommended Lipid-Lowering Therapy After Acute Coronary Syndrome: Opportunities for Improvement. J. Am. Coll. Cardiol. 66, 184–192 (2015).

12. Rodriguez, F. et al. Association of Statin Adherence With Mortality in Patients With Atherosclerotic Cardiovascular Disease. JAMA Cardiol. 4, 206–213 (2019).

13. Karlson, B. W. et al. Variability of low-density lipoprotein cholesterol response with different doses of atorvastatin, rosuvastatin, and simvastatin: results from VOYAGER. Eur. Heart J. - Cardiovasc. Pharmacother. 2, 212–217 (2016).

14. Boekholdt, S. M. et al. Very Low Levels of Atherogenic Lipoproteins and the Risk for Cardiovascular Events: A Meta-Analysis of Statin Trials. J. Am. Coll. Cardiol. 64, 485– 494 (2014).

15. Streja, L., Packard, C. J., Shepherd, J., Cobbe, S. & Ford, I. Factors affecting low-density lipoprotein and high-density lipoprotein cholesterol response to pravastatin in the West Of Scotland Coronary Prevention Study (WOSCOPS). Am. J. Cardiol. 90, 731–736 (2002).

16. Kovanen, P. T., Bilheimer, D. W., Goldstein, J. L., Jaramillo, J. J. & Brown, M. S. Regulatory role for hepatic low density lipoprotein receptors in vivo in the dog. Proc. Natl. Acad. Sci. U. S. A. 78, 1194–1198 (1981).

17. Lagace, T. A. et al. Secreted PCSK9 decreases the number of LDL receptors in hepatocytes and in livers of parabiotic mice. J. Clin. Invest. 116, 2995–3005 (2006).

18. Zhang, D.-W. et al. Binding of Proprotein Convertase Subtilisin/Kexin Type 9 to Epidermal Growth Factor-like Repeat A of Low Density Lipoprotein Receptor Decreases Receptor Recycling and Increases Degradation. J. Biol. Chem. 282, 18602–18612 (2007).

19. Brown, M. & Goldstein, J. A receptor-mediated pathway for cholesterol homeostasis. Science 232, 34–47 (1986).

20. Islam, M. M., Hlushchenko, I. & Pfisterer, S. G. Low-Density Lipoprotein Internalization, Degradation and Receptor Recycling Along Membrane Contact Sites. Front. Cell Dev. Biol. 10, (2022).

21. Pfisterer, S. G. et al. Multiparametric platform for profiling lipid trafficking in human leukocytes. Cell Rep. Methods 2, 100166 (2022).

22. Gaddi, A. et al. Pravastatin in heterozygous familial hypercholesterolemia: Low-density lipoprotein (LDL) cholesterol-lowering effect and LDL receptor activity on skin fibroblasts. Metabolism 40, 1074–1078 (1991).

23. Hagemenas, F. C. & Illingworth, D. R. Cholesterol homeostasis in mononuclear leukocytes from patients with familial hypercholesterolemia treated with lovastatin. Arterioscler. Off. J. Am. Heart Assoc. Inc 9, 355–361 (1989).

24. Hagemenas, F. C., Pappu, A. S. & Illingworth, D. R. The effects of simvastatin on plasma lipoproteins and cholesterol homeostasis in patients with heterozygous familial hypercholesterolaemia. Eur. J. Clin. Invest. 20, 150–157 (1990).

25. Thedrez, A. et al. Homozygous Familial Hypercholesterolemia Patients With Identical Mutations Variably Express the LDLR (Low-Density Lipoprotein Receptor): Implications for the Efficacy of Evolocumab. Arterioscler. Thromb. Vasc. Biol. 38, 592– 598 (2018).

26. Urdal, P., Leren, T. P., Tonstad, S., Lund, P. K. & Ose, L. Flow cytometric measurement of low density lipoprotein receptor activity validated by DNA analysis in diagnosing heterozygous familial hypercholesterolemia. Cytometry 30, 264–268 (1997).

27. Raungaard, B., Heath, F., Brorholt-Petersen, J. U., Jensen, H. K. & Faergeman, O. Flow cytometric assessment of LDL receptor activity in peripheral blood mononuclear cells compared to gene mutation detection in diagnosis of heterozygous familial hypercholesterolemia. Cytometry 36, 52–59 (1999).

28. Powell, E. E. & Kroon, P. A. Low density lipoprotein receptor and 3-hydroxy-3-methylglutaryl coenzyme A reductase gene expression in human mononuclear leukocytes is regulated coordinately and parallels gene expression in human liver. J. Clin. Invest. 93, 2168–2174 (1994).

29. Karlson, B. W., Palmer, M. K., Nicholls, S. J., Lundman, P. & Barter, P. J. Doses of rosuvastatin, atorvastatin and simvastatin that induce equal reductions in LDL-C and non-HDL-C: Results from the VOYAGER meta-analysis. Eur. J. Prev. Cardiol. 23, 744–747 (2016).

30. Adams, S. P., Sekhon, S. S. & Wright, J. M. Rosuvastatin for lowering lipids. Cochrane Database Syst. Rev. (2014) doi:10.1002/14651858.CD010254.pub2.

31. Adams, S. P., Tsang, M. & Wright, J. M. Atorvastatin for lowering lipids. Cochrane Database Syst. Rev. (2015) doi:10.1002/14651858.CD008226.pub3.

32. Reiner, Ž., et al. ESC/EAS Guidelines for the management of dyslipidaemias: The Task Force for the management of dyslipidaemias of the European Society of Cardiology (ESC) and the European Atherosclerosis Society (EAS). Eur. Heart J. 32, 1769–1818 (2011).

33. Nordestgaard, B. G. & Tybjaerg-Hansen, A. IDL, VLDL, chylomicrons and atherosclerosis. Eur. J. Epidemiol. 8 Suppl 1, 92–98 (1992).

34. Varbo, A. et al. Remnant cholesterol as a causal risk factor for ischemic heart disease. J. Am. Coll. Cardiol. 61, 427–436 (2013).

35. Ruuth, M. et al. Susceptibility of low-density lipoprotein particles to aggregate depends on particle lipidome, is modifiable, and associates with future cardiovascular deaths. Eur. Heart J. 39, 2562–2573 (2018).

36. Trinder, M., Francis, G. A. & Brunham, L. R. Association of Monogenic vs Polygenic Hypercholesterolemia With Risk of Atherosclerotic Cardiovascular Disease. JAMA Cardiol. 5, 390–399 (2020).

37. Khera, A. V. et al. Diagnostic Yield of Sequencing Familial Hypercholesterolemia Genes in Severe Hypercholesterolemia. J. Am. Coll. Cardiol. 67, 2578–2589 (2016).

38. Borodulin, K. et al. Cohort Profile: The National FINRISK Study. Int. J. Epidemiol. 47, 696–696i (2018).

39. Goldstein, J. L., Basu, S. K. & Brown, M. S. [19] Receptor-mediated endocytosis of low-density lipoprotein in cultured cells. in Methods in Enzymology vol. 98 241–260 (Academic Press, 1983).

40. Reynolds, G. D. & St. Clair, R. W. A comparative microscopic and biochemical study of the uptake of fluorescent and 125I-labeled lipoproteins by skin fibroblasts, smooth muscle cells, and peritoneal macrophages in culture. Am. J. Pathol. 121, 200–211 (1985).

41. Stephan, Z. & Yurachek, E. Rapid fluorometric assay of LDL receptor activity by DiI-labeled LDL. J. Lipid Res. 34, 325–330 (1993).

42. Spandl, J., White, D. J., Peychl, J. & Thiele, C. Live Cell Multicolor Imaging of Lipid Droplets with a New Dye, LD540. Traffic 10, 1579–1584 (2009).

43. Stirling, D. R. et al. CellProfiler 4: improvements in speed, utility and usability. BMC Bioinformatics 22, 433 (2021).

44. Chang, C. C. et al. Second-generation PLINK: rising to the challenge of larger and richer datasets. GigaScience 4, s13742-015-0047–8 (2015).

45. Mars, N. et al. Systematic comparison of family history and polygenic risk across 24 common diseases. Am. J. Hum. Genet. 109, 2152–2162 (2022).

46. Ripatti, P. et al. Polygenic Hyperlipidemias and Coronary Artery Disease Risk. Circ. Genomic Precis. Med. 13, e002725 (2020).

47. McKinney, W. Data Structures for Statistical Computing in Python. in 56–61 (2010). doi:10.25080/Majora-92bf1922-00a.

48. Harris, C. R. et al. Array programming with NumPy. Nature 585, 357–362 (2020).

49. Virtanen, P., et al. SciPy 1.0: fundamental algorithms for scientific computing in Python. Nat. Methods 17, 261–272 (2020).

50. Hunter, J. D. Matplotlib: A 2D Graphics Environment. Comput. Sci. Eng. 9, 90–95 (2007).

51. Pedregosa, F. et al. Scikit-learn: Machine Learning in Python. J. Mach. Learn. Res. 12, 2825–2830 (2011).

52. Waskom, M. L. seaborn: statistical data visualization. J. Open Source Softw. 6, 3021 (2021).

