## Supplementary Materials for "Linking biological variation to outcomes of statin treatment in the general population"

**Supplementary Methods	1**

Definition of CVD events	1

**Supplementary Tables	2**

Supplementary Table 1	2

**Supplementary Figures	3**

Supplementary Figure 1	3

Supplementary Figure 2	4

Supplementary Figure 3	5

### Supplementary Methods

#### Definition of CVD events

Cardiovascular disease events were defined by the diagnosis codes from the International Statistical Classification of Diseases and Related Health Problems 10th Revision (Version:2019, https://icd.who.int/browse10/2019/en#/).

ICD codes for myocardial infarction: I21, I21.0, I21.1, I21.2, I21.3, I21.4, I21.9, I22, I22.0, I22.1, I22.8, I22.9, I23, I23.0, I23.1, I23.2, I23.3, I23.4, I23.5, I23.6, I23.8, I24.1, 25.2. ICD codes for stroke: I60, I60.0, I60.1, I60.2, I60.3, I60.4, I60.5, I60.6, I60.7, I60.8, I60.9, I61, I61.0, I61.1, I61.2, I61.3, I61.4, I61.5, I61.6, I61.8, I61.9, I63, I63.0, I63.1, I63.2, I63.3, I63.4, I63.5, I63.6, I63.8, I63.9, I64.X.

### Supplementary Tables

#### Supplementary Table 1

Additional medication information for each subject group analyzed. Calculations are shown as percent of all subjects in each group and are based on questionnaire answers which were either 'yes', 'no', or NA - for where the information is missing.

|  |  | Had history of blood pressure medication | | | Taking diabetes medication | | |
| --- | --- | --- | --- | --- | --- | --- | --- |
|  | Subject number | Yes, % | No, % | NA, % | Yes, % | No, % | NA, % |
| CONTROL | 135 | 55 | 6 | 39 | 4 | 10 | 86 |
| STATIN | 133 | 56 | 13 | 33 | 26 | 7 | 68 |
| >35% | 60 | 55 | 8 | 37 | 23 | 5 | 72 |
| >40% | 39 | 62 | 8 | 31 | 26 | 8 | 67 |
| >45% | 30 | 57 | 10 | 33 | 23 | 7 | 70 |

### Supplementary Figures

#### Supplementary Figure 1

A flowchart describing how the final analysis groups and subject numbers were arrived at.

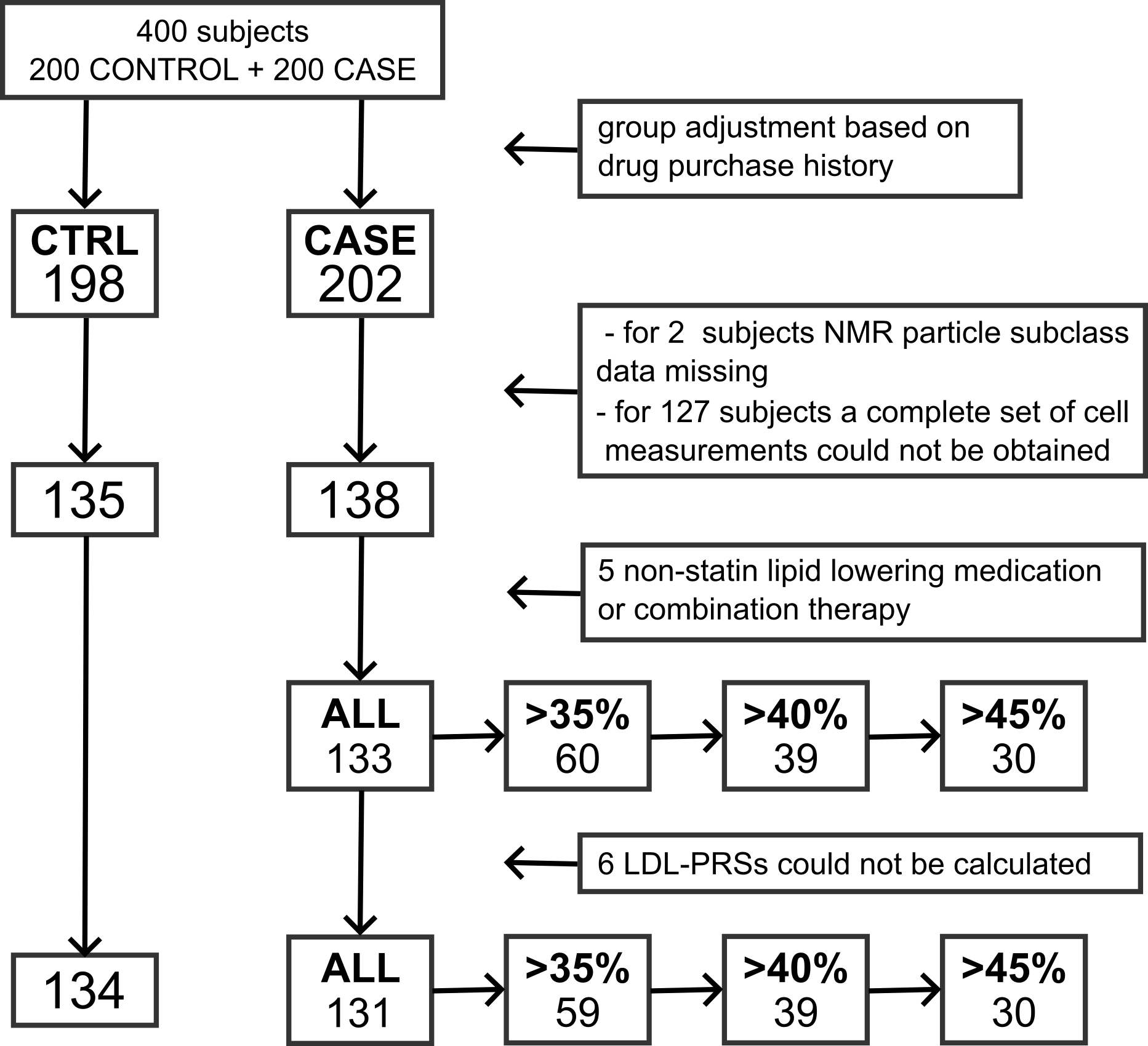

#### Supplementary Figure 2

(A) Correlation of mean LDL-uptake intensity in monocytes under lipid-poor conditions to circulating LDL-C. (B) Correlation of LDL-filled organelle number in monocytes under lipid-rich conditions to circulating LDL-C. (C) Correlation of LDL-filled organelle number in lymphocytes under lipid-poor conditions to circulating LDL-C. (D) Correlation of lipid mobilization in monocytes to circulating LDL-C. Minus sign represents control group (n=135) with no lipid-lowering medication use and plus sign represents subjects on statin monotherapy (n=133). The next three groups are for subjects receiving statin of moderate intensity and higher (>35%, n=60), subjects on high and very high intensity statin (>40%, n=39) and subjects on very high intensity statin therapy (>45%, n=30). * - p<0.05, ** - p<0.01.

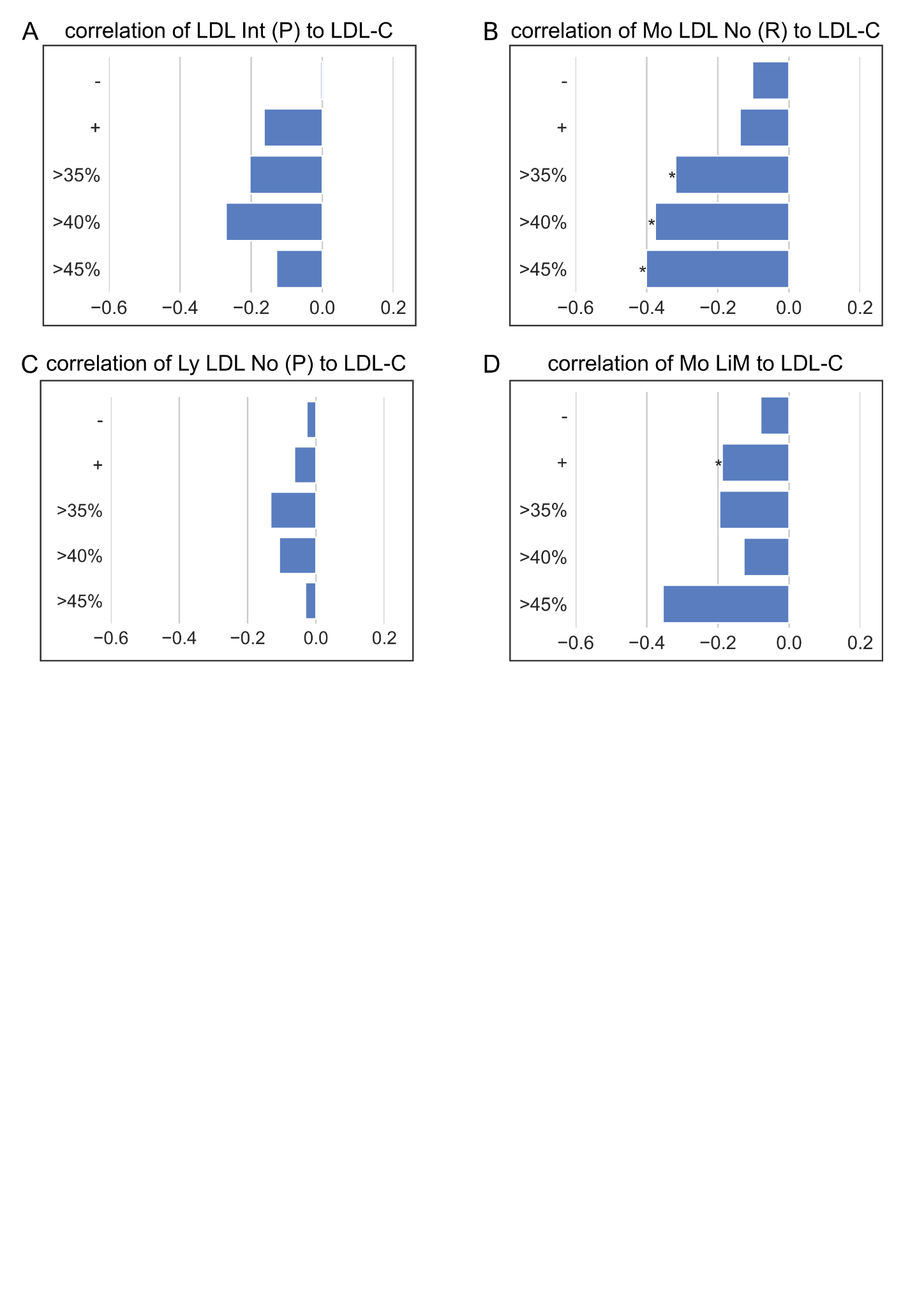

#### Supplementary Figure 3

Spearman's rank correlation between LDL-PRS scores and LDL-C in examined study groups. Minus sign represents control group (n=134) with no lipid-lowering medication use and plus sign represents subjects on statin monotherapy (n=131). The next three groups are for subjects receiving statin of moderate intensity and higher (>35%, n=59), subjects on high and very high intensity statin (>40%, n=39) and subjects on very high intensity statin therapy (>45%, n=30). * - p<0.05, ** - p<0.01.

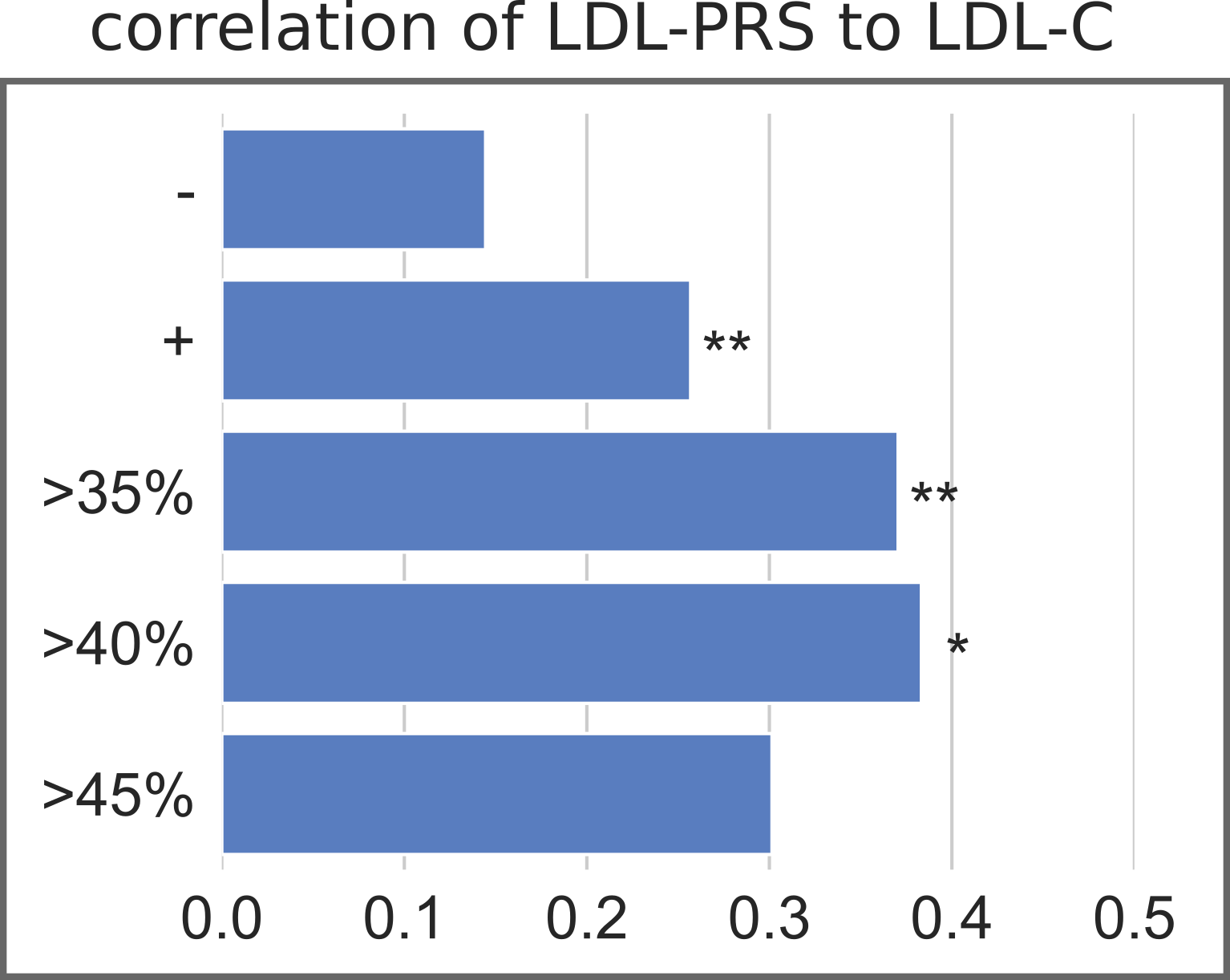
